# Prevalence of COVID-19 and Long COVID in Collegiate Student Athletes from Spring 2020 to Fall 2021: A Retrospective Survey

**DOI:** 10.1101/2022.06.12.22276048

**Authors:** Daisy Massey, Sharon Saydah, Blythe Adamson, Andrew Lincoln, Douglas F. Aukerman, Ethan M. Berke, Robby Sikka, Harlan M. Krumholz

## Abstract

Symptomatic COVID-19 and post-COVID conditions, also referred to as post-acute sequelae of SARS-CoV-2 (PASC) or Long COVID, have been widely reported in young, healthy people, but their prevalence has not yet been determined in student athletes. We surveyed a convenience sample of 18 collegiate school administrators, representing about 7,000 student athletes. According to their survey responses, 9.8% of student athletes tested positive for COVID-19 in spring 2020 and 25.4% tested positive in the academic year of fall 2020 to spring 2021. About 4% of student athletes who tested positive from spring 2020 to spring 2021 developed Long COVID, defined as new, recurring, or ongoing physical or mental health consequences occurring 4 or more weeks after SARS-CoV-2 infection. This study highlights that Long COVID occurs in healthy collegiate athletes and merits a larger study to determine population-wide prevalence.

## Introduction

The longer-term effects of SARS-CoV-2, the virus that causes COVID-19, are only beginning to be explored and understood, but evidence suggests the effects could be substantial. Post-COVID conditions, also referred to as post-acute sequelae of SARS-CoV-2 (PASC), or Long COVID, are defined as new, recurring, or ongoing physical or mental health consequences occurring 4 or more weeks after SARS-CoV-2 infection.^1^ Prevalence of Long COVID based on self-report of symptoms estimates range from 2% to 16%.^2-5^ Long COVID affects all ages and has been reported among persons with severe or mild symptoms from COVID-19.^6^ However, there are fewer estimates of Long COVID among young, healthy adults.^7-9^ Student athletes are a unique group of young, physically active people who can provide insight on the impact of SARS-CoV-2 infection and occurrence of Long COVID. Throughout the pandemic, college athletic departments monitored their athletes closely for both COVID-19 and post-COVID health, including whether or not athletes were able to return to their sport.^10^

This report estimates the prevalence of reported COVID-19, symptomatic COVID-19, and Long COVID in college athletes in the United States attending 18 schools from spring 2020 to fall 2021.

## Methods

We recruited a convenience sample of 18 schools, all members of the National Collegiate Athletic Association (NCAA), the National Junior College Athletic Association (NJCA), and the National Association of Intercollegiate Athletics (NAIA), which responded to the first survey, regarding 2020 spring and the academic year of fall 2020 to spring 2021 (2020-2021). Two schools shared results for 2020-2021 only, due to students not participating in sports in spring 2020. Four of the original 18 schools also responded to the follow-up survey regarding fall 2021.

We developed an online survey to measure the prevalence of student athletes who tested positive for COVID-19, developed Long COVID, and did not return to their sport during the period. The three leagues, which include 1,121 member schools combined, shared contact information for member schools’ healthcare administrators with the research team. The research team distributed an online consent form and survey to healthcare administrators on October 12, 2021.

The survey collected the number of student athletes, COVID-19 cases, cases with Long COVID, and students who had to stop participating in sport for the duration of the relevant time period (spring 2020, 2020-2021, or fall 2021). The survey asked participants to report only cases with a positive COVID-19 test result. We defined Long COVID in accordance with the Centers for Disease Control and Prevention (CDC), which defines post-COVID conditions as symptoms that continue for longer than 4 weeks after infection.^1^ We defined infection after vaccination, in accordance with the CDC, as a case of COVID-19 that occurs after someone has been fully vaccinated (14 days after receipt of a second dose of a mRNA vaccine or receipt of 1 dose of a single-dose vector vaccine).^11^

A follow-up survey was sent to the initial respondents on January 18, 2022, to solicit data from the fall 2021 semester (August 1 to December 30, 2021). Responses were reported in aggregate for each school and no individual student information was provided. This study received exemption from the Yale University IRB. This activity was reviewed by CDC and was conducted consistent with applicable federal law and CDC policy.^§^

Outcomes were reported by school semester or academic year, including spring 2020, 2020-2021, and fall 2021.

## Results

The median number of student athletes per school was 442.5 [95% confidence interval (347.7, 537.3)] in spring 2020 and 454.5 (374.2, 534.8) in 2020-2021. Four of the 18 school administrators shared the number of total student athletes by gender. At these four schools during spring 2020, 55.1% were male, 44.9% were female, and 0% were other gendered (Table 1). During 2020-2021, 52.4% were male, 47.6% were female, and 0% were other gendered. During fall 2021, 50.8% were male, 49.2% were female, and 0% were other gendered. All schools that responded to both surveys were members of the NCAA league. All three NCAA Divisions were represented in the study during spring 2020 and 2020-2021 and, across all time periods, most schools competed in Division I.

**Table 1.** Description and Characteristics of Participating Colleges

|  | 2020<br>Spring<br>Semester | 2020-2021<br>Academic Year<br>(Fall 2020 to<br>Spring 2021) | 2021 Fall<br>Semester |
| --- | --- | --- | --- |
| Overall number of participating schools (N= 1,121) | 16 | 18 | 4 |
| Overall number of eligible student athletes in sample | 6,923 | 7,651 | 1,776 |
| **Schools by Division** |  |  |  |
| Division I | 12 | 13 | 3 |
| Division II | 1 | 1 | 0 |
| Division III | 3 | 4 | 1 |
| **Schools by Region** |  |  |  |
| Northeast | 4 | 5 | 0 |
| Southeast | 6 | 7 | 3 |
| Midwest | 1 | 1 | 0 |
| Southwest | 4 | 4 | 1 |
| Northwest | 1 | 1 | 0 |
| **Schools by Size of Student Athlete Population** |  |  |  |
| Median number of students | 442.5<br>(95% CI<br>347.7,<br>537.3) | 454.5 (95% CI<br>374.2, 534.8) | NA |
| Number of students in 1<sup>st</sup> quartile (smallest programs) | 678 | 1,000 | NA |
| Number of students in 2<sup>nd</sup> quartile | 1,571 | 1,598 | NA |
| Number of students in 3<sup>rd</sup> quartile | 1,998 | 1,957 | NA |
| Number of students in 4<sup>th</sup> quartile (largest programs) | 2,676 | 3,096 | NA |
| **Gender of students (from schools that reported gender among total students)** |  |  |  |
| Total number of schools that reported gender | 4 | 4 | 4 |
| Total number of students with gender reported | 2,060 | 2,145 | 1,776 |
| Male | 1,135 (55.1%) | 1,124 (52.4%) | 902 (50.8%) |
| Female | 925 (44.9%) | 1,021 (47.6%) | 874 (49.2%) |
| Other | 0 | 0 | 0 |

The survey reports represented 6,923 total student athletes in spring 2020 and 7,651 student athletes in 2020-2021. The number of student athletes who tested positive for COVID-19 was 678 (9.8%) in spring 2020 and 1,943 (25.4%) in 2020-2021. Of the student athletes who tested positive, the number of those who were symptomatic was 171 (25.2%) in spring 2020 and 1,082 (55.7%) in 2020-2021. Of the student athletes who tested positive, the number of those who had Long COVID was 29 (4.3%) in spring 2020 and 71 (3.7%) in 2020-2021. Finally, of the student athletes who tested positive, 9 (1.3%) stopped participating in their sport (training or competition) for the rest of the spring 2020 semester and 14 (0.7%) stopped participating in their sport for the rest of the 2020-2021 academic year.

Four schools that had participated in the initial survey also participated in a follow-up survey regarding the fall 2021 semester and representing 1,776 student athletes. For these schools who reported cases from fall 2021, we also compared the percent testing positive, symptomatic, and with Long COVID to rates during the earlier time periods at only these 4 schools. Of the student athletes at these 4 schools, 103 (6.2%) tested positive for COVID-19 in spring 2020, 480 (27.9%) tested positive during 2020-2021, and 406 (22.8%) tested positive in fall 2021. Of the student athletes who tested positive at these schools, 45 (43.7%) were symptomatic in spring 2020, 292 (60.8%) were symptomatic in 2020-2021, and 323 (79.6%) were symptomatic in fall 2021; 4 (3.9%) developed Long COVID in spring 2020, 8 (1.7%) developed Long COVID in 2020-2021, and 4 (1.0%) developed Long COVID in fall 2021; and 0 (0.0%) stopped participating in their sport for the rest of the semester or year in each time period (spring 2020, 2020-2021, and fall 2021).

During 2020-2021, 106 (5.4%) of the 1,943 student athletes who tested positive for COVID-19 had been fully vaccinated against COVID-19 (infection after vaccination), and there was no report of Long COVID among this group.

## Discussion

Our study adds to the understanding of the longer-term effects of COVID-19 in a relatively young, healthy adult population. Our findings indicate that in a young sample population that is especially fit, a significant number of COVID-19 infections can be symptomatic (we found 25.2% to 79.6%, depending on time period) and some people (we found 1.0% to 4.3%, depending on time period) will still experience Long COVID, or symptoms lasting longer than one month. Further study is needed to confirm these estimates of the prevalence of COVID-19, symptomatic COVID-19, and Long COVID among physically active young adults. Given the heterogeneity of Long COVID experiences, determining the prevalence of severe Long COVID among a young healthy population is necessary to target treatment and to educate young people on the risks of COVID-19.^12^

Previous research of young adult populations has indicated that symptomatic infection and Long COVID are less common among the younger adult population.^7-9^ Our results are similar to previous reports. The rate of Long COVID among people ages 18–24 years has been estimated to be 2.2% of people living in private households in the UK, as compared to 2.4% of people all ages.^9^ Regarding athletes, a previous study indicated that test positivity of student athletes in the NCAA was comparable to test positivity of non-athlete students, however the study did not determine the prevalence of COVID-19 or Long COVID within the population.^10^ One study of athletes ages 18–35 years indicated that about half of athletes with COVID-19 were symptomatic 30 days after the initial COVID-19 infection, with 1.2% having moderate or severe symptoms.^13^ Previous research on athletes has shown that Long COVID, in particular cardiac symptoms, can occur and that the physiological impact of COVID-19, including on heart rate, lasts 2-3 months on average.^14-16^

There are several limitations to this study. First, we have a convenience sample of 18 schools that responded to our survey out of 1,121 and only 4 that also responded regarding the fall 2021 follow-up survey. However, these schools are diverse and represent thousands of students, and the survey included all student athletes from each school, which should reduce bias. Additionally, Long COVID symptoms are often ill-defined and underreported, especially during our study period, which could have led to an underreporting of those students who had symptoms more than 4 weeks after initial infection. We were unable to determine the rate of Long COVID among those students who were fully vaccinated and tested positive for COVID-19 due to uneven reporting. Testing and treatment protocols varied between schools and may have affected these comparisons. Finally, characteristics of students, including gender, were not uniformly reported, and were thus excluded from analyses. However, this was a well-monitored population that was under constant surveillance from healthcare administrators.

## Conclusion

Long COVID occurs among young, healthy athletes and is a real consequence of COVID-19. Understanding the prevalence, duration, and lasting consequences of Long COVID requires longer follow-up and further study.

## Data Availability

All deidentified, aggregate data produced in the present study are available upon reasonable request to the authors. Data will be released by league and not by school, to protect student confidentiality.

## Acknowledgments

COVID, Sports, and Society Workgroup

---

**Figure 1.**
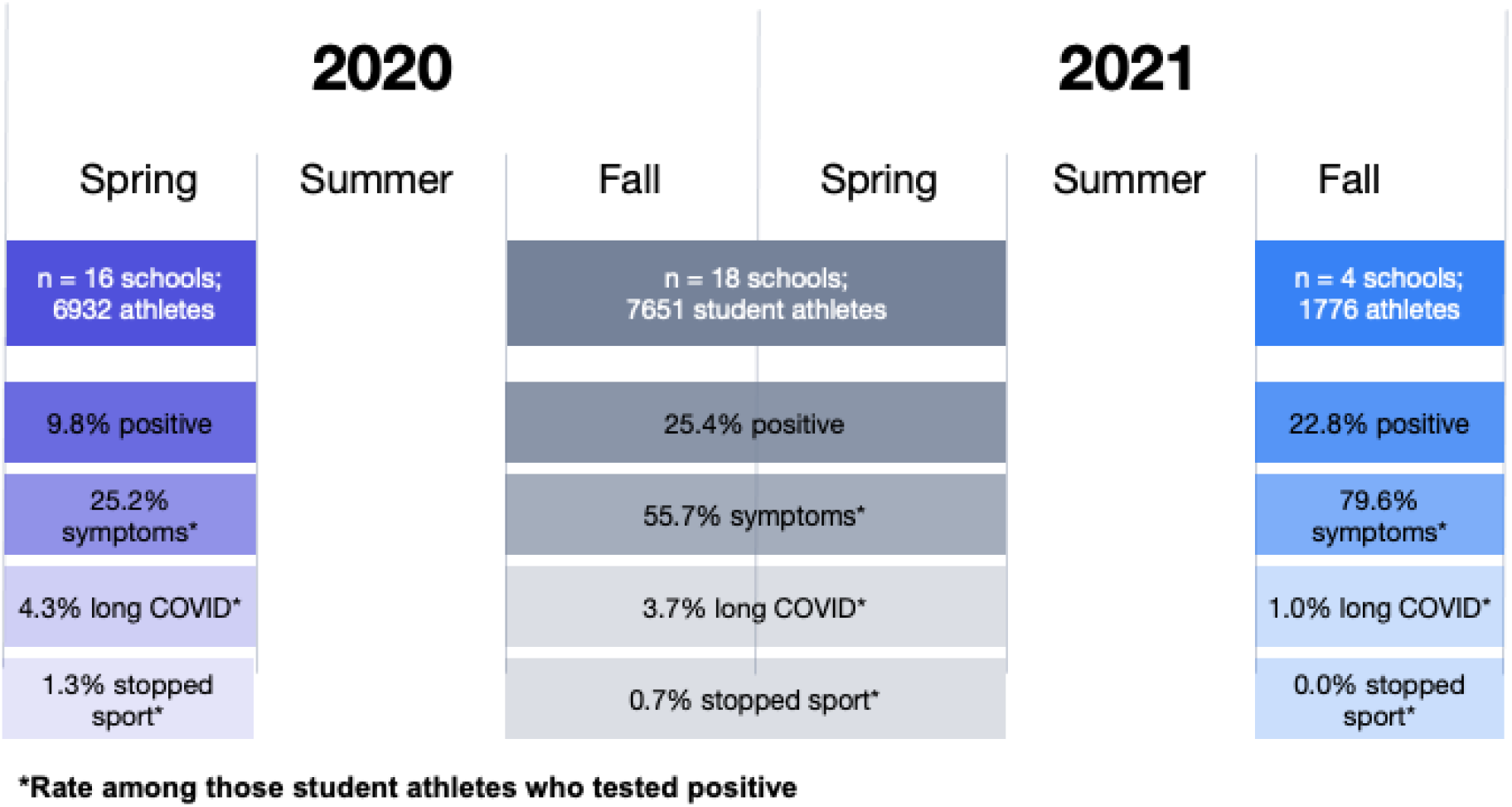
Key findings on the prevalence rate of Long COVID** among COVID-positive student athletes in the NCAA **Long COVID is defined as new, recurring, or on-going physical or mental health consequences occurring 4 or more weeks after SARS-CoV-2 infection.

## Footnotes

§ See e.g., 45 C.F.R. part 46. 102(I)(2), 21 C.F.R. part 56; 42 U.S.C. § 241 (d); 5 U.S.C. §552a; 44 U.S.C. §3501 et seq.

## Notes

### Competing Interest Statement

The authors have declared no competing interest.

### Funding Statement

This study did not receive any funding.

### Author Declarations

This study received exemption from the Yale University IRB. This activity was reviewed by CDC and was conducted consistent with applicable federal law and CDC policy.

